# Beyond Length of Stay: Patient and Carer Perspectives on Virtual Hospital Pathways Following Colorectal Surgery

**DOI:** 10.64898/2026.08.24.26361282

**Authors:** Lillian Reza, Zahra Arabi, Helen Ward, Laura Payne, James Kinross, Vanash Patel

**Affiliations:** Department of Surgery, Anaesthetics and Cancer, West Hertfordshire Teaching Hospitals NHS Trust, Watford General Hospital, Vicarage Road, WD18 0HB, UK; Public Health, Imperial College London, 10th Floor QEQM Building, St Mary’s Hospital, London W2 1NY, UK; Department of Surgery and Cancer, Imperial College London, 10th Floor QEQM Building, St Mary’s Hospital, London W2 1NY, UK

**Keywords:** Virtual Hospital, Patient and Public Involvement, Colorectal surgery, Early discharge

## Abstract

**Background:** Virtual hospital (VH) pathways support early discharge through remote monitoring, but limited evidence has hindered implementation in colorectal surgery. This study aimed to define patient-and carer-relevant outcomes and experiences of VH following colorectal surgery.

**Methodology:** A patient and public involvement and engagement (PPIE) consultation was conducted with 8 participants (7 patients, 1 carer; 4 women, 4 men) who had experienced VH following bowel resection at a high-volume robotic unit. Purposive sampling ensured that 50% of participants had experienced readmission. The 90-minute session was delivered via Microsoft Teams. Data were analysed using reflexive thematic analysis.

**Results:** Seven themes were identified: readmission, remote monitoring, carer burden, recovery, equity, readiness for discharge, and information delivery. Patients supported early discharge when remote monitoring enabled timely detection of complications and readmission pathways were efficient. Readmission was not perceived as failure but as appropriate escalation. Dissatisfaction with readmission was related to delays in emergency care. Remote monitoring provided psychological safety, with patients feeling “held” at home. Carers assumed substantial, often unrecognised, quasi-clinical roles. Recovery was defined by return to function rather than length of stay. Equity concerns were evident, with VH favouring those with adequate support at home, digital literacy, and language proficiency. Discharge readiness was both clinical and psychological. Information delivery at discharge was often poorly retained and requires reinforcement preoperatively at every encounter with patients and carers.

**Conclusions:** VH pathways are acceptable and valued. Readmission is a marker of system responsiveness rather than failure of early discharge on VH. Psychological preparedness, carer support, and equitable access are critical to successful and scalable implementation of early discharge using a virtual hospital.

## BACKGROUND

Virtual hospital (VH) pathways, also described as virtual wards or hospital-at-home (HAH) models, gained prominence during the COVID-19 pandemic in response to unprecedented pressure on inpatient services. Within the National Health Service (NHS), virtual wards using digitally enabled care were subsequently incorporated into the Delivery Plan for Recovering Urgent and Emergency Care Services (2023), reflecting their role in managing ongoing system pressures(1). They have since been identified as a priority in the NHS England 10-Year Plan, supporting a shift towards delivering care closer to home(2).

The VH service at West Hertfordshire Teaching Hospitals NHS Trust (WHTH) was established in 2020 and has since evolved into a mature, integrated model delivering remote monitoring and multidisciplinary care. The service initially focused on patients with COVID-19 and has since evolved to include patients with cardiorespiratory disease and other medical conditions meeting predefined criteria for safe remote management(3). A large-scale propensity-matched cohort study of almost 3,000 patients within this service demonstrated significant reductions in inpatient bed utilisation, readmissions, and healthcare costs. VH reduced length of stay by 3.13 days compared with matched inpatient controls, with total bed-day savings of 13,119 days and a net cost saving of £3.79 million over 33 months of operation(3).

Virtual hospital care offers a natural extension of Enhanced Recovery After Surgery programmes in elective colorectal surgery, a field where adoption of robotic surgical techniques have progressively reduced postoperative pain, inflammatory response, and length of stay(4). Since 2023, the VH service at WHTH has been used to support early discharge in selected patients following colorectal cancer and benign surgery(5). However, evidence supporting the safety, effectiveness, and patient acceptability of VH in surgical populations remains limited. Existing evaluations of VH have largely focused on quantitative endpoints such as length of stay, complication rates, and readmission. In the only randomised controlled trial assessing virtual care with remote monitoring following surgery, readmission was used as a primary marker of pathway success(6,7). While important, these binary metrics may not fully capture the multidimensional nature of postoperative recovery, which encompasses physical, psychological, and social domains.

Clinician-derived outcomes alone are insufficient to evaluate novel care pathways in which patients and carers assume active roles in their own recovery. Patient and public involvement and engagement (PPIE) provides a structured approach to incorporating lived experience into research design and service evaluation, enabling identification of outcome domains that are meaningful to patients rather than defined solely by institutional priorities(8). In the context of VH pathways, where early discharge may shift elements of clinical care into the home environment, understanding the patient and carer perspective is essential to safe and equitable implementation.

This study aimed to explore patient and carer experiences of a VH pathway following colorectal surgery and to define patient-relevant outcome domains to inform future evaluation and interventional research.

## METHODS

### Study Design

We conducted a qualitative PPIE consultation to explore patient and carer experiences of a virtual hospital pathway following colorectal surgery. A focus group methodology was selected to facilitate discussion between participants with shared experience of the pathway, enabling exploration of perspectives and outcomes relevant to patients and carers.

The study was designed and reported in accordance with the GRIPP2 short form reporting checklist, which provides a standardised framework for transparent reporting of patient and public involvement in research(9).

### Participants and Setting

Eight participants were recruited from the VH colorectal discharge pathway at WHTH, including seven patients and one carer. Of the seven patients, four were men. Participants had undergone surgery for colorectal cancer or benign colorectal disease. Purposive sampling was used to ensure diversity of experience, with approximately half of participants having experienced postoperative complications or readmission. The readmission rate in the unit is 7%; purposive over-sampling of this group was intended to capture a range of experiences and identify areas for improvement.

### Data Collection

A 90-minute focus group consultation was conducted via Microsoft Teams in October 2025, facilitated by three researchers with clinical and qualitative research experience (LR, HW, VP). A semi-structured topic guide was used to frame the discussion around four areas: patient and carer experiences of the VH pathway; outcomes considered important by patients and carers; barriers and challenges to early discharge; and the role of community and primary care services following discharge. The session was audio-recorded, transcribed verbatim, and anonymised prior to analysis.

### Data Analysis

Transcripts were analysed using reflexive thematic analysis following the framework described by Braun and Clarke(10). This approach was selected for its flexibility in capturing both explicit accounts and latent meanings within qualitative data, and for its suitability in applied health research contexts where the aim is to identify meaningful outcome domains rather than test predefined hypotheses(11).

Two researchers independently reviewed the transcript and generated initial codes through data analysis. Codes were developed inductively, without reference to a predetermined coding framework. Candidate themes were identified and refined through iterative reflexive discussion between researchers. Final themes were reviewed against the dataset to ensure coherence and completeness.

### Ethical Considerations

This study was conducted as a service evaluation and did not require formal NHS Research Ethics Committee review. All participants were provided with written information about the study prior to the focus group session. Participants provided informed consent to audio recording, and use of anonymised data for research and publication.

## RESULTS

Reflexive thematic analysis of the focus group transcript identified seven interrelated themes organised into four overarching domains that demonstrate how patients and carers perceive safety, recovery, and success within a virtual hospital pathway following colorectal surgery (Figure 1).

**Figure 1:**
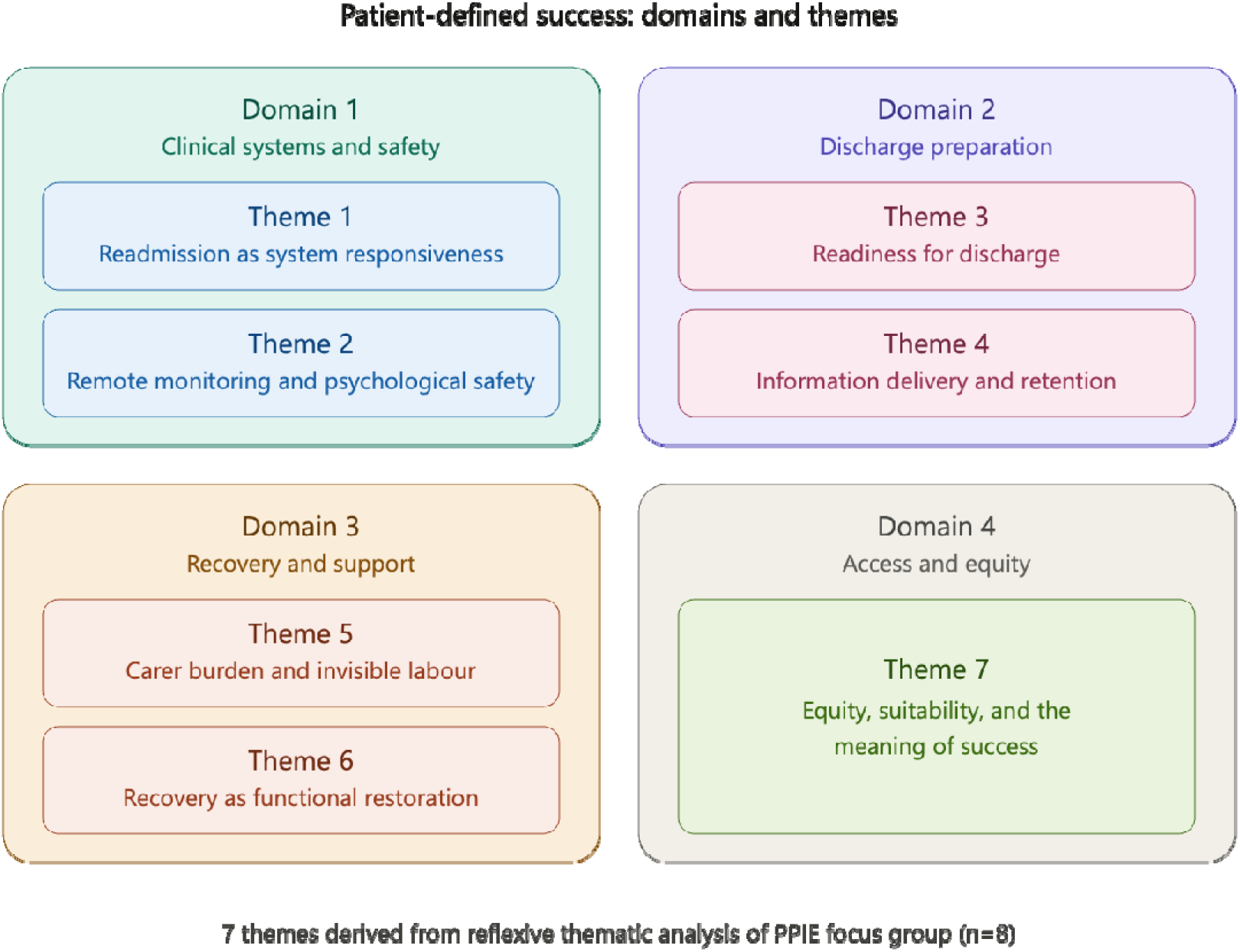
Four domains summarising patient defined success of a VH pathway

### Domain 1: Clinical Systems

#### Theme 1: Readmission as system responsiveness

Readmission was not perceived as a failure of early discharge but as a marker of appropriate clinical escalation and system responsiveness. Participants described readmission as evidence that the pathway was able to detect and manage complications in a timely manner.

> *“The system shouldn’t treat it like you’ve failed recovery; sometimes you need a check or IV antibiotics.”*
>
> *“Could have stayed five days and still had a complication.”*

Participants acknowledged that complications may occur regardless of discharge timing, and that the ability to detect problems early was more important than whether readmission occurred. The ability to return to hospital when required provided psychological safety and was seen as a sign of a clinically responsive team.

Three sub-themes were identified to describe the process of readmission:

- Early detection as success: participants valued the system’s ability to identify complications promptly and considered this to be a positive feature of the pathway.

- Emergency Department as a barrier: the experience of waiting in the Emergency Department (ED) for assessment was identified as the primary point of dissatisfaction associated with readmission. Prolonged waits while acutely unwell could easily change an otherwise positive pathway experience into a distressing one.

> *“If A&E takes twelve hours, that’s the failure, not the readmission.”*

- Direct access pathways: in contrast to long waits in ED, participants who bypassed the ED and were reviewed directly by the surgical team reported positive experiences. Continuity of care with a named consultant or specialist nurse was found to be valuable. Access to the clinical care team varied according to time of day and day of week which was raised as an area for pathway refinement.

The use of readmission as an outcome metric was also concerning for participants when evaluating VH against standard care. Patients managed on standard inpatient pathways may have complications treated during the index admission that are not recorded as readmissions, whereas equivalent events on a VH pathway may result in readmission. This suggests that using readmission rates as an outcome measure to compare systems may disadvantage VH pathways.

#### Theme 2: Remote monitoring and psychological safety

Remote monitoring and regular clinical contact provided reassurance and a sense of being “held” by the clinical team while at home. Participants described the monitoring as an expression of continued care, with monitoring equipment symbolising ongoing hospital presence in the home setting.

> *“Knowing someone was there gave peace of mind.”*

Participants valued contact even when no clinical intervention was required and found that regular ‘check-ins’ reduced anxiety during the recovery period.

Three sub-themes were identified:

- Technology as connection: monitoring equipment was described as simple to use and accessible even for older participants. Monitoring made recovery less stressful and maintained a feeling of connection to the hospital.

- Confidence in self-management: participants found that regular contact increased confidence in being able to manage their own care during the post operative period.

> *“Having contact numbers reduced worry.”*
>
> *“Whatever happens, I can get hold of somebody.”*

- Psychological safety: Feeling monitored and supported was valued independent of clinical need. Participants felt that the sense of being cared for contributed to their recovery and was meaningful in its own right.

### Domain 2: Discharge Preparation

#### Theme 3: Readiness for discharge

Participants described two separate entities required to establish whether they were ready for discharge. Clinical readiness for discharge was defined by objective criteria such as vital signs and mobility. Psychological readiness was defined by confidence, understanding, and emotional security but participants felt that this was not always objectively considered. Discharge decisions were also perceived by some as system-driven rather than patient-centred, with bed availability appearing to take precedence over individual patient readiness. This perception reduced trust in the discharge decision and created anxiety about being discharged prematurely.

> *“It felt like I was discharged because they needed the bed.”*
>
> *“Some people might want to go home early, others aren’t ready — it shouldn’t be one-size-fits-all.”*

Participants identified several criteria they considered important for genuine discharge readiness, including adequate pain control, the ability to mobilise, comprehension of discharge information, carer preparedness, and patient-reported confidence. Weekend discharge was identified as a specific concern due to the possibility of reduced access to clinical support. The presence of a carer at discharge planning and education was considered important by participants.

#### Theme 4: Information delivery and retention

Information provided at discharge was frequently poorly retained. Participants identified two overlapping issues.

- Medication-related cognitive clouding: many participants often received discharge information while still under the effects of postoperative analgesia, significantly impairing their ability to process and retain what they were told.

> *“They explained everything when I was still groggy.”*
>
> *“My partner didn’t know who to call or what was normal.”*
>
> *“I got more from the pre-op nurse than from discharge.”*

- The knowledge gap: participants highlighted that certain complications, such as ileus had not been explained preoperatively, leaving them unable to recognise warning signs during recovery. This created anxiety and, in some cases, delayed escalation.

Participants emphasised the value of repeated and reinforced education delivered at multiple preoperative patient encounters, and not solely at the point of discharge. Carer presence in education was also identified as important, both to share the information burden and to support patients’ with reduced ability to retain information in the immediate postoperative period.

### Domain 3: Recovery and Support

#### Theme 5: Carer burden and invisible labour

Carers assumed substantial quasi-clinical responsibilities with limited formal preparation or support. The single carer participant described the experience as overwhelming as they were taking responsibility for clinical monitoring, medication management, and physical care while also managing competing domestic responsibilities, including the care of a newborn child.

> *“I was taking his blood pressure at home — I’m not a nurse.”*
>
> *“It was stressful because I was caring and working full-time.”*

Carers described a sense of pride in contributing to recovery but felt that the burden of clinical care and support was not obvious to the clinical team.

Participants suggested that carer capacity should be formally assessed as part of the eligibility process for VH discharge, and that this should also consider the impact of caregiving on individuals with other responsibilities such as caring for dependants.

#### Theme 6: Recovery as functional restoration

Participants described recovery as a process of regaining independence and reported ‘feeling recovered’ when they were able to perform daily activities without reliance on carers. Recovery did not correlate with duration of hospital stay or a fixed clinical endpoint.

> *“It wasn’t about days; it was when I could look after myself.”*
>
> *“When I could eat properly again, that’s when I felt recovered.”*

They supported the use of patient-reported functional outcome measures to evaluate the VH pathway against standard care.

### Domain 4: Access and Equity

#### Theme 7: Equity, suitability, and the meaning of success

VH pathways may not be equally accessible or appropriate for all patients. Those with adequate social support, digital literacy, and language proficiency were perceived as better placed to benefit, while those without these resources risked being disadvantaged or excluded.

> *“I had family around; others wouldn’t cope.”*

*“People with poor housing or no support shouldn’t be expected to manage this way.”* Patients who might be otherwise suitable for VH discharge but lacked the social support to manage safely at home should not be excluded. Additional support mechanisms including short term community nursing or home care could be considered for this group. Language barriers and limited technology literacy were also identified as further potential sources of inequity that require pathway support.

Finally, participants suggested that outcomes such as length of stay and readmission rates were not on their own adequate to assess the quality of a VH pathway.

> *“Length of stay makes it sound like a race.”*
>
> *“Readmission isn’t failure; not being listened to is.”*

A successful VH pathway was described as one that made patients feel safe, supported, and confident. Pathway responsiveness to changes in clinical course and continuity of clinician oversight with streamlined escalation were reported as core features for quality assessment in a VH pathway.

## DISCUSSION

Enhanced Recovery After Surgery (ERAS) protocols have transformed perioperative care in colorectal surgery over the past two decades, progressively reducing inpatient length of stay through optimised analgesia and early mobilisation. The adoption of minimally invasive surgical approaches has accelerated this trend further(4,12). Robotic platforms are associated with faster postoperative recovery due to reduced incisional pain through smaller and off-midline extraction sites, and a lower postoperative inflammatory response from reduced tissue injury(5). Despite the advantages of ERAS and robotic surgery, early discharge following colorectal surgery has not been widely accepted and is often treated with caution. Virtual hospital pathways and remote monitoring provide surgeons with a safe paradigm for early discharge in carefully selected patients.

Holtestaul et al. demonstrated in a same-day discharge colorectal pathway, that the barriers to successful early discharge are increasingly non-clinical in nature. Social factors, patient discomfort with discharge, and inadequate carer education prevented same day discharge(13).

This PPIE study explores patient perspectives of a VH pathway following elective colorectal surgery and identifies outcomes relevant to patients for quality assessment. Seven themes were organised into four broad domains. These findings have informed refinement of the surgical pathway and will guide future evaluations of VH pathways against standard care.

A key finding of this study is the reframing of readmission as a marker of system responsiveness rather than pathway failure. In the propensity-matched cohort study from WHTH, unadjusted readmission rates were higher in VH patients than in matched inpatient controls, yet after regression adjustment, rates were significantly lower (3). Readmission following early discharge is not equivalent to readmission following standard inpatient care, and treating these events as comparable may systematically disadvantage VH pathways. Participants prioritised the process of readmission and the timeliness of escalation to the appropriate clinical team.

Remote monitoring and regular clinical contact provided psychological safety and continued connection to the clinical team. This is in keeping with results from the WHTH medical VH cohort, in which anxiety and depression scores improved significantly during the VH admission and 98% of patients reported feeling safe(3). In future evaluations of the VH pathway, psychological outcomes with validated tools for anxiety, confidence in self-management, and patient experience should be assessed.

Information delivery and retention were identified as significant challenges affecting patient experience and engagement with the pathway. Pathway refinement since this study has focused on ensuring that information is delivered and reinforced preoperatively through every clinical encounter with mandatory carer inclusion.

The most significant observation was the impact of VH on the caregiver’s experience. Carers assumed responsibilities that they may not have been fully prepared for and has been so far under recognised. This is not unique to surgical populations; analysis of patients who declined VH care in the WHTH medical service found that 22% cited perceived burden to themselves or caregivers as their reason for refusal of the medical VH pathway(14). Similar findings have been reported in same day discharge pathways. Inadequate education of carers and carer preparedness prevented pathway engagement (13), which further underscores the importance of carer preparedness for pathway success and supports the need for a formal assessment of carer capacity during eligibility assessment for the VH pathway.

VH pathways may preferentially benefit patients with adequate housing, social support, digital literacy, and language proficiency. Without deliberate mitigation strategies, implementation risks widening existing health inequalities. Socioeconomic and domestic circumstance evaluation should form part of eligibility assessment, and enhanced support packages should be available for patients who are clinically eligible but lack the social support for standard VH discharge. The economic impact of this is unclear and necessitates formal evaluation prior to large scale implementation of VH.

This PPIE exploration provides a framework for VH pathway refinement in colorectal surgery and has generated patient-defined outcomes for large-scale evaluation of VH pathways across different healthcare settings.

## Strengths and Limitations

This study provides qualitative insight into patient and carer experiences of a VH pathway following colorectal surgery within a high-volume robotic unit with a mature, integrated VH service. Purposive sampling ensured inclusion of participants with diverse experiences including readmission. The findings are supported by published quantitative data from the same unit, in a larger medical cohort, allowing patient experience to be interpreted alongside clinical outcomes from the same pathway.

The small sample size and single-centre design for VH in colorectal surgery limit generalisability. The findings from this PPIE study however are intended to inform hypothesis generation, outcome selection, and pathway design rather than provide definitive evaluation. A single carer participated, limiting insight into carer experience. The impact of VH on carers and its wider economic implications warrant dedicated qualitative investigation and formal assessment against standard care in future studies.

## DECLARATIONS

### Ethics approval and consent to participate

This study was conducted as a service evaluation within West Hertfordshire Teaching Hospitals NHS Trust and did not require formal NHS Research Ethics Committee approval. All participants were provided with written information prior to participation and gave informed consent for participation, audio recording, and use of anonymised data.

### Consent for publication

All participants provided consent for anonymised data to be used in publication.

### Availability of data and materials

The datasets generated and/or analysed during the current study are not publicly available due to the qualitative nature of the data and the potential risk of participant identification but are available from the corresponding author on reasonable request.

### Authors’ contributions

LR, HW, and VP conceived and designed the study. LR, HW, and VP conducted the focus group and collected the data. LR and ZA performed the data analysis. LR drafted the manuscript. ZA, HW, LP, JK, and VP critically revised the manuscript for important intellectual content. All authors read and approved the final manuscript.

## Data Availability

All data produced in the present study are available upon reasonable request to the authors

